# Detection of SARS CoV-2 contamination in the Operating Room and Birthing Room Setting: Risks to attending health care workers

**DOI:** 10.1101/2021.09.03.21262874

**Authors:** Patricia E. Lee, Robert Kozak, Nasrin Alavi, Hamza Mbareche, Rose C. Kung, Kellie E. Murphy, Darian Perruzza, Stephanie Jarvi, Elsa Salvant, Noor Niyar N. Ladhani, Albert J.M. Yee, Louise-Helene Gagnon, Richard Jenkinson, Grace Y. Liu

## Abstract

**Background:** The exposure risks to front-line health care workers who are in close proximity for prolonged periods of time, caring for COVID-19 patients undergoing surgery or obstetrical delivery is unclear. Understanding of sample types that may harbour virus is important for evaluating risk.

**Objectives:** To determine if SARS-CoV-2 viral RNA from patients with COVID-19 undergoing surgery or obstetrical care is present in: 1) the peritoneal cavity of males and females 2) the female reproductive tract, 3) the environment of the surgery or delivery suite (surgical instruments, equipment used, air or floors) and 4) inside the masks of the attending health care workers.

**Methods:** The presence of SARS-CoV-2 viral RNA in patient, environmental and air samples was identified by real time reverse transcriptase polymerase chain reaction (RT-PCR). Air samples were collected using both active and passive sampling techniques.

**Results:** In this multi-centre observational case series, 32 patients with COVID-19 underwent urgent surgery or obstetrical delivery and 332 patient and environmental samples were collected and analyzed to determine if SARS-CoV-2 RNA was present. SARS-CoV-2 RNA was detected in: 4/24(16.7%) patient samples, 5/60(8.3%) floor, 1/54(1.9%) air, 10/23(43.5%) surgical instruments/equipment, 0/24 cautery filters and 0/143 inner surface of mask samples.

**Conclusions:** While there is evidence of SARS-CoV-2 RNA in the surgical and obstetrical operative environment (6% of samples taken), the finding of no detectable virus inside the masks worn by the medical teams would suggest a low risk of infection for our health care workers using appropriate personal protective equipment (PPE).

## INTRODUCTION

Front line health care providers are at risk of contracting infections when caring for patients with COVID-19^1–3^. Moreover, close, direct and often prolonged patient contact is essential in surgery and obstetrics. However, this may facilitate SARS-CoV-2 infections through the known vectors of respiratory droplets, aerosols and fomites, but infections may potentially be transmitted through exposure to the virus from the surgery or obstetrical delivery itself^4–6^. It remains unclear if the type of surgical/obstetrical procedure, may present different risks to HCWs attending these patients.

SARS-CoV-2, the virus that causes COVID-19 can spread via respiratory droplets and aerosols of infected persons through coughing, sneezing or talking^7–10^. Additionally, the virus has been documented to be present in the gastrointestinal (GI) tract and consequently any surgery that involves opening the GI tract (bowel related surgery) is thought to pose a risk to medical teams^11,12^. There are case reports of SARS-CoV-2 virus detected in peritoneal fluid from a patient with COVID-19 undergoing surgery^13,14^. In the female reproductive tract, SARS-CoV-2 RNA has been identified in amniotic fluid, vaginal swabbing and documented cases of vertical transmission have been found^15–18^. Potentially, if the virus is present on peritoneal surfaces of males or females, in the female reproductive tract or the myometrium, this virus could be aerosolized via cautery smoke, or, from the release of CO_2_ gas from laparoscopic procedures. While there is no current published research on the presence of SARS-CoV-2 in the surgical smoke/plume, there is existing literature which identified other viruses including human papillomavirus (HPV), Human Immunodeficiency Virus −1 and Hepatitis B virus in surgical smoke^19–28^.

The risks of aerosolization from the respiratory tract is recognized^29^, but the risk of SARS-CoV-2 residing in the surgical site and the subsequent risk of aerosolizing this virus from the surgical site is not well studied. SARS-CoV-2 has been found in air samples with research demonstrating that the virus contained in aerosol particles can remain viable in the air for extended periods of time and are potentially infectious after both human shedding and airborne transport^30,31^. There is documentation of SARS-CoV-2 virus in the hospital ward setting^6,32–34^, in the delivery suite^35^ and with tracheostomies (an aerosol generating medical procedure)^29^.

More importantly, reports on the risks of SARS-CoV-2 viral contamination of the HCW in the operating room (OR) and birthing room setting (apart from studies on tracheostomies) were not identified by this author; although there have been studies reporting on HCW mask viral contamination in the patient ward setting^36–39^. Since a respiratory route is considered the main route of infection, knowledge of the risks of contamination of the masks is critical to assess the potential risks of viral exposure to the HCW working in the OR/delivery suites.

As COVID-19 is likely to continue to circulate and be endemic, even in the presence of vaccines, knowledge of risks will be essential to determine the optimal means to protect HCWs.

We seek to study the risk of contamination in the OR and birthing suite environment via evaluating the risk of aerosolization from the respiratory tract or from the surgical/obstetrical field during surgery or labor and delivery. This information is key to assessing the risks to HCWs who care for patients at the time of 1) vaginal delivery or cesarean section and 2) other surgical procedures whether they are vaginal, abdominal or laparoscopic. This knowledge would help guide best practice regarding the use of personal protective equipment (PPEs) and safety in the OR and birthing room.

The objectives of this study are to determine if SARS-CoV-2 viral RNA from patients with COVID-19 undergoing surgery or obstetrical care is present in: 1) the peritoneal cavity (males/females), 2) female reproductive tract, 3) on surgical instruments/equipment used, 4) the floor of the procedure rooms, 5) bioaerosols produced during surgery or obstetrical delivery, and 6) inside surgical masks of the attending health care workers.

## METHODS

### Study Design

From November 2020 to May 2021, patients with a nasopharyngeal (NP)/mid-turbinate (MT) swab positive for SARS-CoV-2 by RT-PCR (reverse transcriptase polymerase chain reaction), in need of urgent surgery or obstetrical delivery at one of two large academic Toronto hospitals: Sunnybrook Health Sciences Centre (Sunnybrook) or Sinai Health System, were prospectively identified by the surgical/obstetrical clinical teams. This study was approved by both hospitals’ Research Ethics Boards. Environmental samples (air samples and floor samples) in the operating room or delivery suite were obtained with no patient or HCW consent required.

Following informed consent, patient samples (peritoneal fluid, vaginal, myometrial or placental swabs) were collected and following HCW consent, samples were obtained from their masks. Mask sampling was offered to any HCW in the room caring for the patient. Depending on the type of surgical case, patient consent was not possible, not always sought, or, not relevant to the study samples collected. Consenting HCWs agreed to follow up with hospital Occupational Health if SARS-CoV-2 RNA was detected on their mask.

Patient clinical and laboratory data were obtained by chart review and/or participant interview.

### Eligibility

Patients included those known to test positive by NP swab either symptomatic or asymptomatic within 30 days of diagnosis; or known symptomatic COVID-19 positive patients beyond 30 days of diagnosis. HCWs included any consenting HCW present in the operating/delivery room, caring for the patient. Study Samples (Appendix A included for detail):

Patient sampling for any laparotomy cases included peritoneal cavity fluid (male or female). Patient samples for obstetrical cases included: vaginal fluid and swabs of the myometrium (at time of cesarean section) and membranous placenta.

Equipment and environmental samples included: swabs of the room floor (within 1 metre from the surgical site, and 2 metres away from the surgical site with a 10×10cm area for 17 cases, and a 30×30cm area for the final 13 cases^6^; collection of the cautery filter; swab of equipment (e.g. endotracheal tube, saw blade, surgical instruments etc); and swab of the inside of the surgical mask worn by HCWs^37,40,41^. Swabbed samples were collected with either a sterile flocked swab or sterile dental pledget that was pre-moistened with universal transport media (UTM) and immediately placed in 3cc of UTM.

Bioaerosol sampling was obtained via two previously described methods: 1) Active air sampling via the GilAir Plus sampler was used at two locations: as close to the surgical site as possible (within 0.5-1 metres) and 2-3 metres away (Sensidyne®, https://www.sensidyne.com/air-sampling-equipment/gilian-air-sampling-pumps/gilair-plus/)^42^. Samples were collected with a 37 mm three-piece cassette with 0·8 μm polycarbonate filter, sampling at a rate of 3.5L/min for the duration of the procedure with PCR detection after elution from the filter 2) Passive air sampling was considered in the last 10 cases and involved an open Petri dish to collect any viral particles settling by gravity in the dish (within 1-2 metres of the patient)^34,35,40,43,44^.

### Laboratory Methods

All collected samples were processed at the Shared Hospital Laboratory (Sunnybrook). Aside from the cautery and active air sample filters, the lab staff were blinded to the source of the sample.

Virus detection was performed by real-time RT-PCR using a multi-target assay currently utilized in the laboratory for detection of the virus^45^.

Additionally, the Ct (cycle threshold) value of the assay as an estimate of the viral load was used and where possible was obtained from the initial diagnostic swab of the patient.

The outcome of interest was SARS-CoV-2 RNA PCR positive samples. Whole genome sequencing was not performed with our surgical/obstetrical samples, but was performed on diagnostic nasopharyngeal swabs when possible, in order to identify variants of concern (VOC).

#### Sample Size

The primary outcome was the rate of SARS-CoV-2 RNA PCR positive samples from the HCWs masks. The expected outcome was 0% positivity. We planned to study 40 patients with an expected mean of 2-3 HCW masks per patient (or 100 mask samples) which, with an expected positive rate of 0%, would provide a 95% confidence interval range of 0-5% (R Statistical Software: R version 3.5.3, 2019). The study was closed after sampling from 32 patients, with the collection of 143 masks which provided adequate data to study the primary outcome. An interim analysis after the first 20 patients recruited was performed and based upon new published studies, a decision was made to add passive air sampling to the protocol^34,35,40,43^.

Descriptive statistics, Shapiro-Wilk normality test, Mann-Whitney U test and 2-sample test for equality of proportions with continuity correction tests were used where appropriate (R Statistical Software: R version 3.5.3, 2019).

## RESULTS

### Patients

A total of 32 patients with COVID-19 (Table I), 18 female and 14 male were enrolled (mean 53.55 years, SD = 18.068, range: 20-88 years): 9 obstetrical patients (5 cesarean sections, 4 vaginal births) from Sunnybrook and Sinai Health System, and 23 semi/urgent surgical patients from Sunnybrook’s Divisions of General Surgery (7/23), Trauma/Orthopedic Surgery (7/23), Gynecologic Oncology (2/23), Burn (2/23), Plastic Surgery (1/23), Cardiac Surgery (1/23), Vascular Surgery (1/23), Gastroenterology (1/23) and Neurosurgery (1/23).

**Table 1:** Patient and case characteristics with results of samples taken.

| ID # | Age range (yrs) | Sex | No. days from 1<sup>st</sup> COVID-19 positive test to surgery (repeat test) | Surgical Division | Procedure performed | VOC | NP swab (Ct value)<br>a, b | Positive <sup>c</sup> samples obtained (Ct value) | Active air sampling time (min) | Negative <sup>d</sup> samples | No. negative HCW masks <sup>e</sup> |
| --- | --- | --- | --- | --- | --- | --- | --- | --- | --- | --- | --- |
| 1 | 30-39 | F | 3 | OB | Caesarean section (spinal anesthesia) | Not tested | - (n/a) | - (none) | 74 | •peritoneal fluid, placenta, myometrium, •floor | 3 |
| 2 | 30-39 | F | 4 | OB | Caesarean section (spinal anesthesia) | Not tested | - | - | 137 | •peritoneal fluid, placenta, myometrium, •floor | 3 |
| 3 | 40-49 | F | 3 | OB | Caesarean section (spinal anesthesia) | Not tested | 30.89 | •placenta (25.9) | 89 | •vaginal fluid<br>•patient mask<br>•floor | 5 |
| 4 | 20-29 | F | 30 (24) | OB | Caesarean section (intubation and extubation) | Alpha | 18.69 | •ETT (25.66) | 125 | •peritoneal fluid, placenta, myometrium, •floor | 6 |
| 5 | 20-29 | F | 14 (0) | OB | Caesarean section (spinal anesthesia) | Not VOC | 34.29 | - | Not sampled | •peritoneal fluid, placenta | 4 |
| 6 | 30-39 | F | 3 | OB | Caesarean section | Alpha | 24.41 | •Vaginal fluid (23.45)<br>•Peritoneal fluid (25.1)<br>•myometrium (28.34) | Not sampled | •floor | 3 |
| 7 | 30-39 | F | 1 | OB | Vaginal delivery | Not tested | 31.31 | - | Not sampled | • vaginal fluid<br>•patient mask<br>•floor | 3 |
| 8 | 30-39 | F | 0 | OB | Vaginal delivery | Not tested | 18.87 | - | Not sampled | •vaginal fluid, placenta, •floor | 1 |
| 9 | 30-39 | F | 14 (0) | OB | Vaginal (vacuum) Delivery | Alpha | 14.81 | - | Not sampled | •vaginal fluid<br>•placenta<br>•integrated visor of mask (obstetrician) | 6 |
| 10 | 50-59 | F | 1 | Gyne Oncology | TAH BSO, node dissection (intubation and extubation) | Not tested | 32.17 | - | 146 | •ETT<br>•floor | 5 |
| 11 | 40-49 | F | 16 | Gyne Oncology | TAHBSO omentectomy (intubation and extubation) | Alpha | 11.86 | •ETT (25.37)<br>•surgical clamps, scissors (27.13)<br>•floor at 2-metres (28.94) | 149 | •floor near the OR table | 6 |
| 12 | 80-89 | F | 4 | Ortho | L hip hemiarthroplasty (spinal anesthesia) | Not tested | 21.44 | - | Not sampled | •saw blade<br>•floor | 1 |
| 13 | 30-39 | F | 2 | Ortho | Bilat tibial fracture (spinal anesthesia) | Not tested | - | - | 176 | •drill bit<br>•floor | 5 |
| 14 | 50-59 | M | 22 (3) | Ortho | T10-T12 spinal decompression (arrived in OR intubated and left intubated) | Not tested | 29.17 | •ETT (22.27) | 242 | •floor | 6 |
| 15 | 70-79 | M | 2 (0) | Ortho | ORIF cervical spine fracture (arrived in OR intubated and left intubated) | Not tested | - | - | 211 | •Petri dish<br>•floor | 5 |
| 16 | 60-69 | F | 4(2) | Ortho | ORIF ankle, fibula | Alpha | 32.35 | - | 106 | •scalpel blade and clamps<br>•Petri dish<br>•floor | 5 |
| 17 | 50-59 | M | 2 | Ortho | Humerus fracture (came to OR intubated and left OR intubated) | Not VOC | 13.45 | •floor (21.97) | 97 | •scissors and clamps<br>•Petri dish | 5 |
| 18 | 70-79 | M | 26 (5) | Ortho | Superficial and deep compartment irrigation of leg and debridement (arrived in OR intubated and left OR intubated) | Alpha | 37.25 | - | 68 | •scalpel blade, scissors and clamps<br>•Petri dish<br>•floor | 4 |
| 19 | 50-59 | M | 1 | General Surgery | Laparotomy (came to OR intubated and left intubated) | Not tested | 32.7 | - | 189 | •Peritoneal fluid,<br>•floor<br>•face shield (surgeon) | 4 |
| 20 | 50-59 | F | 15 | General Surgery | Bilat mastectomy (intubation and extubation) | Not tested | - | - | 123 | •ETT<br>•floor | 5 |
| 21 | 70-79 | M | 70 (3) | General Surgery | Tracheostomy | Not tested | - | •ETT (21.74) | 111 | •floor | 5 |
| 22 | 60-69 | M | 50 (5) | General Surgery | Tracheostomy | Alpha | - | - | 69 | •ETT<br>•floor | 5 |
| 23 | 60-69 | M | 54 (16) | General Surgery | Tracheostomy | Not VOC | - | •ETT (22.53)<br>•floor (28.19) | 68 | •floor at 2 metres | 4 |
| 24 | 70-79 | F | 62 (2) | General Surgery | Tracheostomy, gastroscopy, insertion of gastrostomy tube | Not VOC | 32.94 | •ETT (20.7)<br>•floor (20.9) | 117 | •clamps and scissors<br>•Petri dish | 5 |
| 25 | 70-79 | M | 4 | GI | Gastro duodenoscopy | Alpha | 18.62 | •Gastroscope (30.56) | 62 | •Petri dish<br>•floor | 3 |
| 26 | 60-69 | F | 2 | Burn | Burn reconstruction (intubation and extubation) | Not tested | - | •ETT (24.88) | 142 | •floor<br>•face shield (surgeon) | 7 |
| 27 | 70-79 | M | 2 | Burn | Debridement, graft lower abdomen, thighs (intubation and extubation) | Beta/ Gamma | 26.69 | •ETT (24.32)<br>•floor (24.97)<br>•Petri dish UTM (28.4) | 173 | •scalpel blade, scissors and clamps | 4 |
| 28 | 20-29 | M | 2 | Plastics | Hand surgery (regional block) | Not tested | 35.29 | - | 136 | •floor | 5 |
| 29 | 60-69 | F | 1 | Plastics | Debridement and graft (arrived in OR intubated and left intubated) | Alpha | 36.01 | - | 217 | •dermatome •floor | 6 |
| 30 | 60-69 | M | 14 | Cardiac Surgery | Aortic valve replacement (not extubated) | Not VOC | 32.98 | - | 285 | •thoracic retractor<br>•floor | 6 |
| 31 | 70-79 | M | 2 | Vascular Surgery | Carotid endarterectomy (intubation and extubation) | Not tested | - | - | 178 | •ETT<br>•floor<br>•face shield (surgeon) | 4 |
| 32 | 40-49 | M | 0 | Neuro Surgery | Decompressive Craniotomy (arrived in OR intubated and left intubated) | Not tested | - | - | 77 | •floor | 4 |

\*Patient consent was obtained
Yrs, years; VOC, variant of concern; NP, nasopharyngeal; Ct, cycle threshold: the number of cycles required for the fluorescent signal to cross the threshold in RT-PCR (a lower Ct would indicate a higher viral load); HCW, health care worker; M, male; F, female; OR, operating room; bilat, bilateral; ETT, endotracheal tube; n/a, not available; OB, Obstetrics; Ortho, Orthopedic Surgery; TAHBSO, total abdominal hysterectomy bilateral salpingo-oophorectomy; Alpha, Alpha variant (United Kingdom, B.1.1.7); ORIF, open reduction internal fixation; GI, gastrointestinal; Beta/Gamma, Beta or Gamma variant (South African, B.1.351 or Brazilian, P.1), UTM, universal transport medium
<sup>a</sup> The mean Ct value of the initial patient NP swab was 21.72, 95% CI 16.90-27.29 and 29.96 95% CI 27.07-33.43 respectively in the group of patients that had positive samples (evidence of SARS-CoV-2 RNA) and the group that had all samples negative (Mann-Whitney U test, p=0.007).
<sup>b</sup> The mean Ct value of the initial patient NP swab was 24.52 (95% CI: 18.72-30.99) and 28.41 (95% CI: 25.35-30.33) respectively in the group of patients that were known VOC and the group that included known non-VOC and unknown/untested status (Mann-Whitney U test, p=0.4561).
<sup>c</sup> Positive samples refer to the samples collected (patient, instrument, equipment, surface, air and mask) that have the presence of SARS-CoV-2 RNA detected on real time RT-PCR (reverse transcriptase polymerase chain reaction)
<sup>d</sup> Negative samples refer to the samples collected (patient, instrument, equipment, surface, air and mask) that do not have the presence of SARS-CoV-2 RNA detected on real time RT-PCR
<sup>e</sup> A variety of masks were used by HCWs in our cohort including American Society for Testing and Materials (ASTM) level 3 masks, N95 masks, double masks (inner mask sampled) or masks worn under face shields. Of 143 masks: 51 from nurses, 31 from anesthetists, 32 from surgeons and 29 from surgical house staff.

Of the 32 patients enrolled, the patient’s first SARS-CoV-2 positive nasopharyngeal swab occurred a median of 4 days before their procedure (mean 13.77, range: 0-70 days). 11/32 patients had a repeat NP swab closer to the date of their procedure (median 3 days, mean 5.08, range: 0-24 days).

### Samples tested

A total of 343 samples were taken for SARS-CoV-2 RNA detection: 11/343 were duplications (1 patient submitted 2 masks; 10/12 endotracheal tubes were sampled twice with different methodology: flocked swab (4/12 samples positive for viral RNA) and dental pledget (7/10 samples positive). Twenty of the 332 (6.02%) samples tested positive for SARS-CoV-2 RNA (Table II): 8/12 endotracheal tubes (3/10 tested positive with both flocked swab and dental pledget), 1/6 peritoneal fluid (1/5 cesarean section cases, 0/1 trauma laparotomy case), 1/7 placentas, 1/4 myometrial swabs, 1/7 vaginal fluid, 2/11 samples from surgical equipment of 11 different surgical cases (positives found from the scissors and clamps from an abdominal hysterectomy and from the gastroscope used intra-operatively for a gastroduodenoscopy), 1/7 passive air samples, and 5/60 floor samples (5/24 of the 30×30cm samples and 0/36 of the 10×10cm samples were positive for SARS-CoV-2 RNA: 4/30 samples taken 1 metre and 1/30 samples taken 2 metres from the surgical field were positive). There were no positive samples for SARS-CoV-2 RNA from among cautery filters (0/24 cases); active air sampling (0/47, 25 cases) and the inside of HCWs masks (0/143, 32 cases sampled). 0/2 patient masks sampled were positive.

**Table II:** Positive tests for SARS-CoV-2 RNA (number of positive samples/number of patients or cases sampled)

| **Surgical Service**<br>**(No. of cases)** | **Peritoneal fluid <sup>a</sup>** | **Vaginal fluid <sup>a</sup>** | **Myometria l swab <sup>a</sup>** | **Placenta swab <sup>a</sup>**<br>**(fetal side)** | **Surgical instruments <sup>a</sup>** | **ETT <sup>a</sup>** | **Cautery filter <sup>a</sup>** | **Air sample from GilAir pump filter<sup>b</sup>** | **Petri dish UTM (passive air sampling)<sup>a</sup>** | **Floor swab<sup>b</sup>** | **Face shield<sup>a</sup>** | **Masks <sup>a</sup>** |
| --- | --- | --- | --- | --- | --- | --- | --- | --- | --- | --- | --- | --- |
| Obstetrics<br>N=9 | 1/5 | 1/7 | 1/4 | 1/7 | -- | 1/1 | 0/4 | 0/5 | -- | 0/14 | 0/1 | 0/34 |
| Gynecology<br>Oncology<br>N=2 | -- | -- | -- | -- | 1/ 1 | 1/ 2 | 0/2 | 0/3 | -- | 1/4 | -- | 0/11 |
| Orthopedics/<br>Trauma<br>N=7 | -- | -- | -- | -- | 0/5 | 1/ 1 | 0/6 | 0/11 | 0/4 | 1/14 | -- | 0/31 |
| General<br>Surgery<br>N=2 | 0/1 | -- | -- | -- | -- | 0/1 | 0/2 | 0/4 | -- | 0/4 | 0/1 | 0/9 |
| Tracheo-<br>stomy<br>N= 4 | -- | -- | -- | -- | 0/1 | 3/ 4 | 0/4 | 0/8 | 0/1 | 2/8 | -- | 0/19 |
| Gastro-<br>enterology<br>N=1 | -- | -- | -- | -- | 1/1 | -- | -- | 0/2 | 0/1 | 0/2 | -- | 0/3 |
| Burn/Plastic<br>Surgery<br>N=3 | -- | -- | -- | -- | 0/2 | 2/2 | 0/3 | 0/8 | 1/1 | 1/8 | 0/1 | 0/22 |
| Cardiac/<br>Vascular<br>Surgery<br>N=2 | -- | -- | -- | -- | 0/1 | 0/1 | 0/2 | 0/4 | -- | 0/4 | 0/1 | 0/10 |
| Neuro<br>Surgery<br>N=1 | -- | -- | -- | -- | -- | -- | 0/1 | 0/2 | -- | 0/2 | -- | 0/4 |
| **Total: 32 patients** | **1/6** | **1/7** | **1/4** | **1/7** | **2/11** | **8/12** | **0/24** | **0/47** | **1/7** | **5/60** | **0/4** | **0/143** |

-- (empty cell), sample was either: not taken, not applicable or patient consent was not obtained; ETT, endotracheal tube; UTM, universal transport medium; Plastics, Plastic Surgery;
<sup>a</sup> number of positive tests/number of cases sampled
<sup>b</sup> number of positive tests/number of tests performed (1-2 tests per surgical/obstetrical case)

In 5 surgical cases, the initial positive diagnostic test was beyond 30 days: 1 cesarean section (30 days) and 4 tracheostomies (50, 54, 62 and 70 days). Four of 5 of these cases (30, 54, 62, and 70 days) had positive endotracheal tube samples, and 2 of the 4 endotracheal tube positive cases had positive floor samples (54 and 62 days since diagnosis of COVID-19).

Variants of concern (VOC) were identified in 10 cases. Of the 10 VOC cases: 9 were the Alpha variant (United Kingdom/B.1.1.7), and 1 was either Beta or Gamma variant (South African/B.1.351 or Brazilian/P.1). Of the 10 VOC cases, the following sites tested SARS-CoV-2 RNA positive: 3/4 endotracheal tubes (75%), 2/18 floor samples (including one sample 2 metres away), 1/5 passive air samples, 2/6 cases where the surgical instruments were tested. In addition, the positive peritoneal fluid, myometrial swab and vaginal fluid swab samples came from the same patient who tested positive for the Alpha VOC. There was no significant difference between the proportion of positive samples when comparing the VOC group (n=10) versus unknown/not VOC group (n=22).

The cycle threshold (Ct) value of the initial NP swab positive for SARS-CoV-2 RNA was recorded for 20/32 patients (median: 30.3, mean: 26.65, range: 11.86–37.25). For the VOC group, 9/10 had Ct values recorded (median: 24.41, mean: 24.52 [95% CI: 18.72-30.99], range: 11.86–37.25). For the group that is unknown/not VOC, 11/22 have Ct values recorded (median: 31.10, mean: 28.41 [CI: 25.35-30.33], range: 13.45–35.29). There was no significant difference in Ct values between these two groups (Mann-Whitney U test, p=0.4561). The Ct values of the initial NP swabs were significantly lower (indicating higher viral loads) in those tested who were subsequently found to have any study sample with SARS-CoV-2 RNA (13/32: Ct recorded in 8/13) versus those without any positive study samples (19/32: Ct recorded in 12/19) with a mean Ct 21.72: 95% CI 16.90-27.29 vs mean 29.96: 95% CI 27.07-33.43 respectively (Mann-Whitney U test, p=0.007).

## DISCUSSION

Several studies have documented potential risks of SARS-CoV-2 infection to HCWs in the clinic or hospital ward settings^1,6,29,46^. To our knowledge, this is the first study evaluating potential exposure risks to HCWs in the operating room with a variety of surgical procedures. The OR is a unique environment as HCWs are in prolonged and very close contact (hands on) with patients. In our study, we detected SARS-CoV-2 RNA in non-respiratory patient samples (peritoneal fluid, vaginal fluid, myometrium, placenta), surgical equipment/instruments (endotracheal tubes, gastroscope, laparotomy surgical clamps & scissors), and the surgical room environment (floor, air); but no contamination of the surgical masks worn by HCWs was detected.

Our study corroborates earlier studies that have shown evidence of virus in the respiratory tract and in some of the surgical/obstetrical fields^12,13,15–18,47–49^. We have documented evidence of SARS-CoV-2 RNA in the GI tract, peritoneal cavity, and female genital tract, all of which could potentially be sources of aerosolized virus/viral particles. We did not find evidence of viral RNA in the orthopedic equipment sampled (e.g. saw blade and drill bits), retractors used in cardiac/thoracic surgery and the dermatome used in burn surgery. This may indicate that SARS-CoV-2 does not reside in this type of tissue or at least not present with a viral load high enough for detection.

With our study and others reporting the finding of virus in the peritoneal cavity laparoscopy (and its positive insufflation pressure) theoretically may be considered an aerosol generating procedure justifying the use of appropriate PPE and best practice measures^11,13,14,50–54^.

We used standard techniques for air and floor sampling and found evidence of aerosolization of SARS-CoV-2^6,34,35,40,42^. While the frequency of positive tests was low, this does indicate that aerosolization of the virus does occur in surgery. It is possible that the true positive rates are higher since there are known limitations with the sampling and testing techniques used in this study^6,34,35,40,42^.

We looked for characteristics of the patients’ infections that would increase the risks of detection of viral RNA in the surgical/obstetrical fields or local environment. Higher viral load detected on the initial NP swab (as estimated by the Ct threshold) was associated with a higher risk of detectable virus in our samples, while the subtype of the SARS-CoV-2 virus was not (although the numbers were insufficient for sub-analysis on viral subtyping).

This study was not able to determine if the origin of aerosolized/droplet virus is arising from the surgical fields in the smoke plume. Others did not detect SARS-CoV-2 in electrocautery smoke despite using high viral loads in an *in vitro* setting^28^. While the lack of any positive viral RNA found on the smoke evacuator filters tested would indicate that the viral contamination from the surgical field is absent or below detection limits, these results cannot be used to definitely conclude that surgical smoke does not harbour SARS-CoV-2.

Since infection with SARS-CoV-2 is primarily via the respiratory tract, we chose to sample the inside of HCWs masks as a means of identifying viral contamination in close proximity to the HCWs respiratory tract. Face mask sampling has previously been shown to be an efficacious way of detecting *Mycobacterium tuberculosis* contamination^55^ and has been used to detect SARS-CoV-2 contamination of masks worn by HCW exposed to COVID-19 infected patients on wards (0/25 positive, inside surface) and directly from patient masks (6/10 positive)^37^. Others have studied SARS-CoV-2 viral contamination on the outer surface of face shields worn by HCWs attending women with COVID-19 in labor (one vaginal delivery with all face shields tested being positive)^35^. We sampled the inside of masks and found 0/143 HCWs masks and 0/4 HCWs’ face shields to be positive for SARS-CoV-2 RNA. Our study did not detect SARS-CoV-2 RNA on the inner surface of any mask used by HCWs. This is reassuring, since this finding indicates a low risk of HCWs involved with surgery or obstetrical care being exposed to SARS-CoV-2 RNA if using appropriate PPE.

### Limitations

Our study did not include data on the involved HCWs to determine information regarding previous infections or vaccinations, or if they developed subsequent COVID-19 infections.

Detection of SARS-CoV-2 RNA in bioaerosols is recognized to be challenging, being highly dependent upon air flow, exchange rates and source of emissions (reviewed by Borges et al) and it is suggested that parallel sampling with more than one technique may increase sensitivity^56^. Thus, we employed two air sampling techniques, active air sampling used previously by two of our coauthors (finding 3/146 positive air samples taken from patient rooms)^42^ and the passive technique similar to what is described by others for SARS-CoV-2 virus^34,35,40,43^.

Despite these efforts, we recognize that it is likely that not all viral contamination was detected with this study. Further, even though viral RNA was detected in some samples, this study did not determine if infectious virus was present.

## CONCLUSION

There is evidence of SARS-CoV-2 RNA in the surgical and obstetrical patient’s operative environment (surgical surfaces and aerosolized). However, the finding of no detectable virus on the inner surface of masks worn by the health care teams reassuringly suggests a low risk of infection when wearing appropriate personal protective equipment.

## Supporting information

Appendix A (eMethods)

## Data Availability

The data is displayed in the manuscript's Tables I and II.

## ACKNOWLEDGEMENTS

We are grateful to our operating room nurses and our obstetrical nurses who participated in this study as well as the patients, their anesthetists and their surgeons.

