## Appendix A (eMethods) for "Detection of SARS CoV-2 contamination in the Operating Room and Birthing Room Setting: Risks to attending health care workers"

For Sample Collection:

GENERAL DETAILS:

Each dental pledget was pre-moistened with up to 3cc of sterile UTM from a newly opened unused 15cc sterile Falcon tube containing 3 cc of UTM, universal transport medium (UTM®, Copan Diagnostics, CA, USA; https://www.copanusa.com/sample-collection-transport-processing/utm-viral-transport/).

Hand hygiene and glove changes were performed between the collection of every sample and the outside surface of each container was wiped off with a CaviWipes™ towelette and then each sample was stored in a separate new biohazard marked ziploc plastic bag and placed in the fridge at 4° Celsius within 20 minutes. All collected samples were processed at the Shared Hospital laboratory, located at the Sunnybrook site (stored in the interim at -20° and then -80° Celsius).

Sinai Health samples were stored in the fridge locally and transferred to Sunnybrook within 24 hours.

Samples were analyzed by RT-PCR as previously described (Vermeiren C, Marchand-Senecal X, Sheldrake E, et al. Comparison of Copan Eswab and FLOQswab for COVID-19 PCR diagnosis: working around a supply shortage. J Clin Microbiol 2020.)

All the operating rooms (including those in the Birthing area) have 20 air exchanges an hour.

Cycle threshold values (the number of cycles required for the fluorescent signal to cross the threshold in RT-PCR) quantified viral load, with lower values indicative of a higher viral load.

PATIENT SAMPLES

1) PERITONEAL SAMPLE:

Taken by a member of the obstetrical/surgical team: 5-10 cc of peritoneal fluid (if present upon entering the cavity), or, if no fluid seen, 10cc of sterile saline was placed in the peritoneal cavity with a 10cc sterile syringe and then whatever volume of fluid aspirated back into the syringe was placed in an 80cc sterile plastic container.

2) VAGINA:

Taken by a member of the obstetrical team: a vaginal speculum was placed in the vagina before delivery (typically after informed consent and well before active labor, or, before cesarean delivery) and up to 5 cc of pooled vaginal fluid was aspirated with a sterile 10cc syringe. If no vaginal fluid was seen, 10cc of sterile saline was placed in the vagina with a 10cc sterile syringe and then whatever volume of fluid that was aspirated back into the syringe was placed in an 80cc sterile plastic container.

3) MYOMETRIUM:

Taken by a member of the obstetrical team at the time of cesarean delivery, after the baby is delivered and hemostasis managed, a flocked swab (iClean, HCY, Shenzhen, China; https://www.chenyanglobal.com/oropharyngeal-nylon-flocked-swab-product/) was used to wipe the incised surface of the myometrium and the swab placed immediately in a 15cc sterile plastic Falcon tube containing 3cc of UTM (UTM®, Copan Diagnostics, CA, USA; https://www.copanusa.com/sample-collection-transport-processing/utm-viral-transport/).

4) PLACENTA:

Taken by a member of the obstetrical/research team at the end of the case with a flocked swab which was used to wipe the surface of the membranous placenta and the swab was placed immediately in a 15cc sterile plastic Falcon tube containing 3cc of UTM.

ENVIROMENTAL SAMPLES

5) FLOOR:

Taken by a member of the research team at the end of the case: a sterile dental pledget (3/8" x 1.5" cylindrical sponge, SDP Inc. Montreal, Canada; https://www.sdpmedical.com/en/cylindrical-sponges) was pre-moistened with universal transport media and the floor was swabbed in a location as close to the patient as permits and also 2 metres away. The swabbing was performed in a standardized fashion over a 10x10cm area with 2 perpendicular "S" swipes (or over a 30x30cm area for the last 12 of 32 cases). The pledget was placed immediately in a 15cc sterile plastic Falcon tube containing 3cc of UTM.

6) ENDOTRACHEAL TUBE (ETT):

Taken by a member of the research team at the end of the case after the patient was extubated: a sterile dental pledget or a flocked swab was pre-moistened with UTM and used to wipe the length of the distal half of the ETT and the pledget/swab placed immediately in a 15cc sterile plastic Falcon tube containing 3cc of UTM.

7) SURGICAL INSTRUMENTS / EQUIPMENT:

Taken by a member of the research team at the end of the case: a sterile dental pledget was pre-moistened with UTM and used to wipe the part of the instrument or equipment that was in direct contact with the patient's surgical site. The pledget then placed immediately in a 15cc sterile plastic Falcon tube containing 3cc of UTM.

8) PASSIVE AIR SAMPLE:

3cc of sterile UTM was placed in a sterile Petri dish was placed open by a member of the research team at the beginning of the case within 1-2 metres of the patient and in a location that would not interfere with patient care, on a Mayo stand about 1 metre high from the floor. The Petri dish was retrieved at the end of the case and the UTM in it transferred to a sterile Falcon tube.

9) ACTIVE AIR SAMPLE:

A member of the research team set up 2 air samplers at the start of the case: one in a location that was as close as possible to the surgical site (within 1 metre) and without interfering with patient care; and a second one in a location that was 2-3 metres away from the surgical site. The GilAIr pump was turned on when the patient was brought into the OR and the pump was stopped when the patient left the room. The Polytetrafluoroethylene (PC) filter cassette containing sampled air was wiped on its outer casing with a CaviWipes™ towelette (https://www.metrex.com/en-ca/caviwipes) and placed in a new biohazard marked ziploc bag.

For obstetrical patients with non-operative delivery, one air sampler was placed within 1 metre of the patient's head and at the level of her mouth; the second sampler was placed approximately 2 metres away from the perineum.

10) CAUTERY FILTER:

If cautery was used, at the end of the case, a member of the research team removed the cautery filter device (marVac grey box, https://www.klsmartin.com/en/products/electrosurgery/smoke-evacuation/) and its surface was wiped with a CaviWipes™ towelette and the filter housed in its original box was placed in a new biohazard marked ziploc bag and then placed in the freezer.

11) MASK SAMPLE:

Masks were swabbed on their inside surface by a member of the research team with a sterile dental pledget that was pre-moistened with UTM – wiping over the inside of the mask twice in the area (up to 10x10cm) that would have been in contact with the nose and mouth. The pledget then placed immediately in a 15cc sterile Falcon tube containing 3cc of UTM.

12) FACE SHIELD:

The outer facing surface of the face shield (at least a surface area of 10 x 15 cm) was wiped with a sterile flocked swab that was pre-moistened with sterile UTM and the swab then placed immediately in a 15cc sterile Falcon tube containing 3cc of UTM.
